# Peripheral inflammatory markers relate to central inflammation and survival in syndromes associated with frontotemporal lobar degeneration

**DOI:** 10.1101/2024.01.31.24302080

**Authors:** Maura Malpetti, Peter Swann, Kamen A Tsvetanov, Leonidas Chouliaras, Alexandra Strauss, Tanatswa Chikaura, Alexander G Murley, Nicholas Ashton, Peter Barker, P Simon Jones, Tim D. Fryer, Young T. Hong, Thomas E Cope, George Savulich, Duncan Street, W Richard Bevan-Jones, Timothy Rittman, Kaj Blennow, Henrik Zetterberg, Franklin I. Aigbirhio, John T. O’Brien, James B. Rowe

**Author notes:** Corresponding Author: Dr. Maura Malpetti, Department of Clinical Neurosciences University of Cambridge Herchel Smith Building, Forvie Site, Robinson Way, Cambridge Biomedical Campus Cambridge□CB2 0SZ. Joint Senior authors (John T. O’Brien, James B. Rowe).

## Abstract

Neuroinflammation is an important pathogenic mechanism in many neurodegenerative diseases, including those caused by frontotemporal lobar degeneration (FTLD). There is a pressing need for scalable and mechanistically relevant blood markers of inflammation to facilitate drug development and experimental medicine. We assessed inflammatory profiles of serum cytokines from 214 patients with FTLD-associated syndromes (behavioural and language variants of frontotemporal dementia, progressive supranuclear palsy, corticobasal syndrome). We tested the association with brain microglial activation (by positron emission tomography) and survival. A pro-inflammatory profile across the FTLD spectrum (including TNF-α, TNF-R1, M-CSF, IL-17A, IL-12, IP-10 and IL-6) differentiated patients (all syndromes) from controls. A higher pro-inflammatory profile scores was associated with higher microglial activation in frontal and brainstem regions, and with lower survival. Blood-based markers of inflammation could increase the scalability and access to neuroinflammatory assessment of people with dementia, to facilitate clinical trials and experimental medicine studies.

## Introduction

The pathologies of frontotemporal lobar degeneration (FTLD) cause a diverse family of clinical disorders ^1^, including the behavioural variant of frontotemporal dementia (bvFTD) ^2^, the non-fluent (nfvPPA) and semantic variants of primary progressive aphasia (svPPA) ^3^, right temporal variant frontotemporal dementia ^4^, progressive supranuclear palsy (PSP) ^5^ and corticobasal syndrome (CBS) ^6^. These disorders vary in their cognitive and motor features, and molecular pathology correlates, but share a pressing need for new therapeutic strategies.

These disorders also have in common evidence for neuroinflammation as a pathogenic process ^7^, from preclinical ^8^, post mortem ^9–13^ and genome wide association studies ^14–16^. The neuroinflammation manifests as microglial and astroglial activation, and increased secretion of inflammatory markers, including tumour necrosis factor (TNF) and interleukin (IL) cytokines ^7^. Systemic autoimmune diseases are more prevalent in patients with FTLD-related disorders than in healthy controls or people with Alzheimer’s disease ^17,18^. Immunomodulatory and anti-inflammatory treatment strategies have been proposed to slow or prevent disease progression. However, a better understanding of the inflammatory profiles and scalable *in vivo* markers are required to inform individual prognosis, disease progression models and trial design.

One can localise and quantify neuroinflammation *in vivo* using positron emission tomography (PET). PET ligands for the mitochondrial translocator protein (TSPO), such as [^11^C]PK11195, have been implemented as *in vivo* proxies of activated microglia, showing increased cerebral inflammation in patients across all of the FTLD-syndromes ^19–24^. TSPO PET has shown that higher inflammation at baseline strongly correlates with clinical severity and predicts faster clinical decline over the following years in FTD, PSP and Alzheimer’s disease ^25–27^.

It follows that immunotherapeutic strategies might reduce the risk and progression of clinical phases of FTLD-related syndromes. However, a barrier in therapeutic development and clinical trials is the lack of scalable and repeatable assays of clinically relevant neuroimmune signals. PET imaging is neither readily scalable, due to cost and radiation dosing, nor easily accessible for many centres and countries. PET also only allows one to test a single molecular target at a time rather than capturing a comprehensive set of inflammatory markers. Fluidic markers can overcome these limitations, providing more scalable biomarkers and insight on the interaction between central and peripheral inflammatory processes.

Few clinical studies have evaluated peripheral blood markers of inflammation in patients with FTLD-syndromes and their progression. Although blood measures do not necessarily capture the level or distribution of brain inflammation, they may nonetheless reflect risk or activity of microglia-mediated and other inflammatory cascades of relevance to experimental medicine studies. They could also enable the formulation of a more complete disease framework, to assist prognosis, stratification and monitoring in trials, and identification of novel therapeutic targets. Importantly, clinically relevant and mechanistically informative blood markers for inflammation would also make research more accessible to people who cannot access to major facilities or are too impaired to take part into neuroimaging-only research studies. Biochemical rather than cell-based assays would also have the advantage of simpler handling and storage for multicentre studies. However, such applications of biochemical markers of inflammation would require evidence of their level and pattern in relation to multiple disorders, and evidence of a significant relationship to cerebral inflammation.

The aim of this study was to identify clinically relevant patterns of blood-based inflammatory cytokines in people with clinical syndromes associated with FTLD. Specifically, we applied a combination of data- and hypothesis-driven approaches to analyse levels of serum cytokines and their associations with survival. We tested the secondary hypothesis that peripheral blood cytokines correlated with inflammation of the central nervous system, as indexed by [^11^C]PK11195 PET.

## Results

Two hundred and fourteen (n=214) patients with clinical diagnoses associated with FTLD and twenty-nine (n=29) cognitively unimpaired controls were recruited Fifty-two patients met diagnostic criteria for bvFTD ^2^, 51 for primary progressive aphasia, comprising 31 cases of nfvPPA, and 20 of svPPA ^3^, 58 for PSP ^5^ and 53 for CBS ^6^. Patients were age- and sex-matched with controls (Table 1). As expected, patients with bvFTD were on average younger than patients with nfPPA, PSP and CBS. Controls had higher ACE-R scores and lower CBI-R scores than patient groups. Patients with PSP had higher ACE-R scores than patients with bvFTD and svPPA. Controls had lower CBI-R scores than all patient groups, except for nfPPA.

**Table 1.** Demographic and clinical characteristics for control and patient groups (*= significant group effect on variance, by 1-way ANOVA).

| **Group** | **N** | **Sex**<br>**F/M** | **Age (years)**<br>**Mean ± SD** | **ACE-R (/100)**<br>**Mean ± SD** | **CBI-R (/150)**<br>**Mean ± SD** |
| --- | --- | --- | --- | --- | --- |
| **HC** | 29 | 13/16 | 68.2 ± 7.9 | 96.2 ± 2.7 | 9.5 ± 8.7 |
| **bvFTD** | 52 | 22/30 | 63.5 ± 9.3 | 64.7 ± 19.2 | 83.1 ± 31.6 |
| **svPPA** | 20 | 7/13 | 67.6 ± 4.8 | 61.6 ± 18.7 | 74.5 ± 34.1 |
| **nfPPA** | 31 | 20/11 | 71.6 ± 7.7 | 69.5 ± 21.8 | 32.9 ± 28.0 |
| **PSP** | 58 | 29/29 | 69.5 ± 6.3 | 79.9 ± 12 | 49.8 ± 29.6 |
| **CBS** | 53 | 27/26 | 70.3 ± 7.5 | 73.9 ± 17.9 | 53.5 ± 33.9 |
| **Group Difference (F)** | | n.s. | 6.3, $p < 0.001^*$<br>(n.s. controls vs patient groups) | 16.4, $p < 0.001^*$<br>(HC > all patient groups) | 22.5, $p < 0.001^*$<br>(HC < all patient groups, except nfPPA) |

*Abbreviations: bvFTD=behavioural variant frontotemporal dementia; nfPPA=non-fluent primary progressive aphasia; svPPA=semantic variant primary progressive aphasia; PSP= progressive supranuclear palsy; CBS= corticobasal syndrome; ACE-R=revised Addenbrooke's Cognitive Examination; CBI-R= Cambridge Behavioural Inventory Revised; F=Female; M=Male; SD= standard deviation; n.s.= non-significant*

### Group differences in the *pattern* of cytokine values

Group-based dissimilarity analyses across 25 cytokines and the related hierarchical cluster complete-linkage analysis identified greater distance values between controls and each patient group than between diagnostic groups (Supplementary Figure 1). The smallest distance value was found between PSP & CBS groups and between bvFTD & CBS groups. The largest distance between patient groups were found between svPPA & nfPPA. In other words, the relative distribution of cytokine concentrations – not the magnitude of their concentration – differentiated all patient groups with an FTLD-disorder from controls.

### Cytokine-derived principal components (cytokine profiles)

The highly significant Bartlett’s test indicated that the cytokine data was suitable for principal component analysis (PCA; Chi-square=856.6, p<0.00001). The Kaiser-Meyer-Olkin (KMO=0.79) criterion also indicated the suitability for PCA. Applying the three criteria described in “Methods”, the first two components were selected for further analyses. Component 1 and Component 2 explained 21.5% and 11.0% of the variance, respectively. Component 1 was positively loaded mainly onto TNF-α, TNF-R1, M-CSF, IL-17A, IL-12, IP-10 and IL-6, while Component 2 was negatively weighted mainly by MCP-4, MCP-1, Eotaxin and TARC (see Figure 1A for contribution of each cytokine).

**Figure 1.**
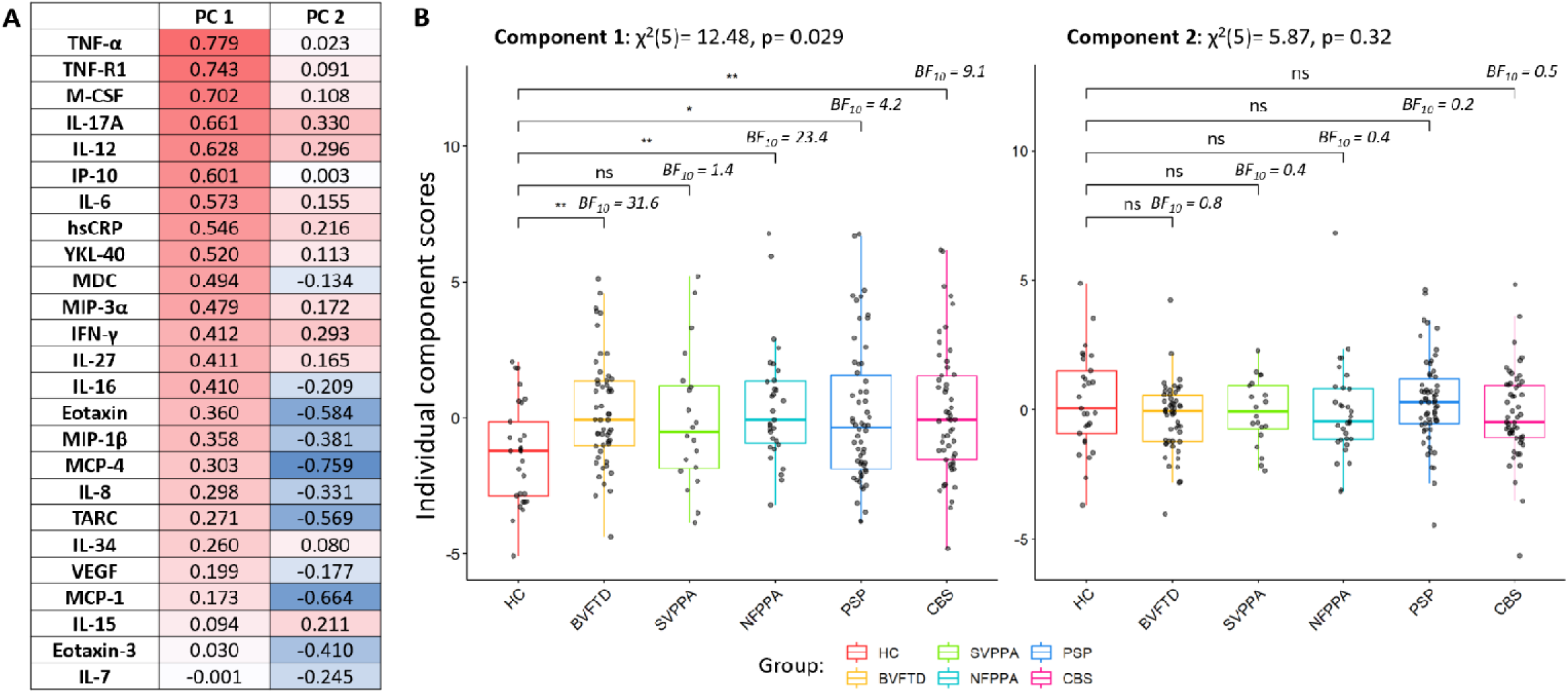
Cytokine-derived principal components (PC) and group comparisons. Panel A: contribution of each cytokine to component 1 and component 2; red colours represent positive correlations, while blue negative correlations. Panel B: group comparisons assessed by Kruskal-Wallis one-way analysis and Dunn’s post hoc tests (P value ***=0.001, **=0.01, *=0.05, BF10=Bayes Factor in favour of the alternate hypothesis and against the null; HC=healthy controls, AD=Alzheimer’s dementia, MCI=mild cognitive impairment, DLB=dementia with Lewy bodies, bvFTD=behavioural variant of frontotemporal dementia, PPA=primary progressive aphasia, PSP=progressive supranuclear palsy, CBS=corticobasal syndrome, MND=motor neurone disorder).

### Patient groups show elevated inflammation

Kruskal-Wallis one-way analysis of variance on Component 1 identified significant group differences (12.48, p =0.029). Dunn’s Multiple Comparison post hoc test highlighted significant differences between controls and each diagnostic group (controls vs bvFTD: 3.12, p=0.002; nfPPA: 3.03, p=0.003; PSP: 2.32, p=0.021; CBS: 2.78, p=0.005), except for patients with svPPA (1.72, p=0.086) (see Figure 1B, and Supplementary Table 2 for pairwise comparisons). Kruskal-Wallis one-way analysis on Component 2 did not identify any significant group differences (Figure 1B). Bayesian pairwise post hoc tests confirmed the results obtained with the frequentist approach (see Figure 1B for BF).

### Inflammation is associated with neurodegeneration biomarkers

The peripheral blood cytokine profile correlated with biomarkers of neurodegeneration. Specifically, the univariate correlations between cytokine-derived Component 1 individual scores and plasma markers identified weak but positive associations with plasma levels of NfL (rho=0.113, p= 0.06, BF=0.7), GFAP (r=0.159, p= 0.015, BF=3.2) and pTau217 (r=0.145, p= 0.025, BF=3.9), across all patients (see Supplementary Figure 2). Multiple regression models with age, sex and diagnosis as covariates confirmed the association between cytokine profile Component 1 and NfL (F=4.48, p=0.036), GFAP (F=4.69, p=0.032) and pTau217 (F=4.08, p=0.045). Bayesian correlation results highlighted the weakness of the evidence for these associations (1 < BF < 3). Univariate correlations and regression models did not identify statistically significant associations between cytokine-derived Component 2 individual scores and plasma markers (p > 0.05).

### Higher inflammation indicates worse prognosis

We tested Component 1 as a prognostic biomarker (in patients only) using Cox proportional hazards regression using age, sex, disease groups as covariates, and cytokine Component 1 and days from blood test to death as predictors of interest for survival. The individual participant loadings on Component 1 were significantly associated with time to death [hazard ratio 1.1 (1.02–1.19), p=0.013] (see Table 2, *“Basic Model”*). To illustrate this effect, we plotted separately the patients with high and low values on this component, as separated by the median (Figure 2). When adding baseline plasma NfL, symptom duration and ACE-R scores to the model as proxies of disease severity, the predictive value of Component 1 on survival remained significant [hazard ratio 1.16 (1.04–1.30), p=0.007] (Table 2, *“Model with severity”)*.

**Figure 2.**
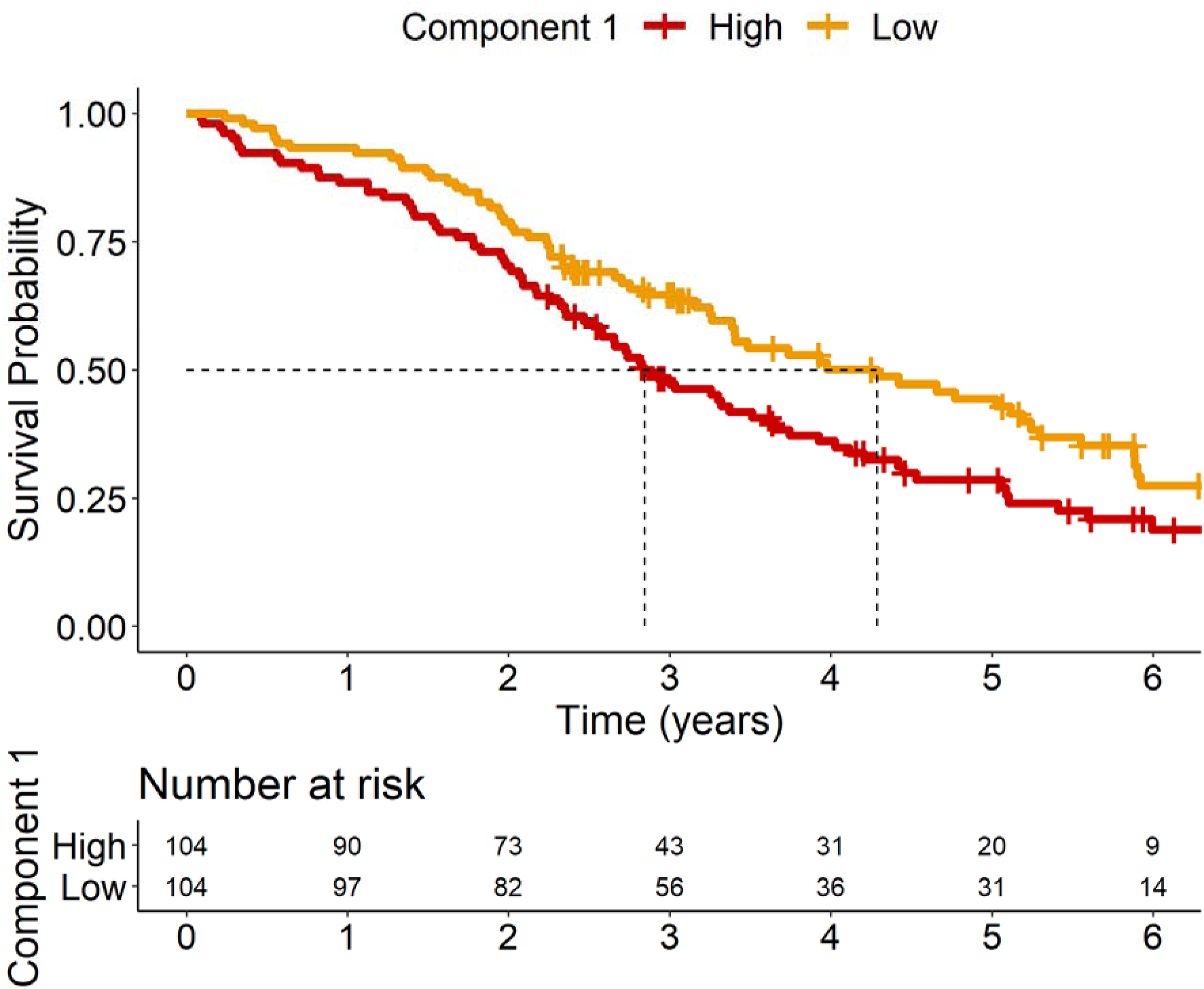
Kaplan–Meir Survival Curve of loadings on Component 1. Patients were separated into two groups based on their loading onto Component 1, with high loadings being greater than or equal to median (≥ −0.191) and low loadings being lower than the median (< −0.191).

**Table 2.** Cox proportional hazards model of time from blood sample to death.

| **Basic Model (n=208)** |  | **HR** | **CI Lower**<br>**95%** | **CI Upper**<br>**95%** | **SE** | **Z** | **Pr(>\|z\|)** |
| --- | --- | --- | --- | --- | --- | --- | --- |
| **Component 1** |  | **1.103** | **1.021** | **1.192** | **0.039** | **2.486** | **0.013** |
| Age |  | 1.053 | 1.025 | 1.082 | 0.014 | 3.815 | <0.001 |
| Sex | Female (ref) |  |  |  |  |  |  |
|  | Male | 1.322 | 0.935 | 1.870 | 0.177 | 1.579 | 0.114 |
| Diagnosis | BVFTD (ref) |  |  |  |  |  |  |
|  | CBS | 1.384 | 0.825 | 2.324 | 0.264 | 1.230 | 0.219 |
|  | NFPPA | 0.552 | 0.291 | 1.047 | 0.327 | -1.818 | 0.069 |
|  | PSP | 1.499 | 0.902 | 2.490 | 0.259 | 1.563 | 0.118 |
|  | SVPPA | 0.484 | 0.242 | 0.967 | 0.353 | -2.055 | 0.040 |
| **Model with Severity (n=133)** |  | **HR** | **CI Lower**<br>**95%** | **CI Upper**<br>**95%** | **SE** | **Z** | **Pr(>\|z\|)** |
| **Component 1** | **PC1** | **1.176** | **1.052** | **1.315** | **0.057** | **2.851** | **0.004** |
| Age | Age | 1.061 | 1.015 | 1.108 | 0.022 | 2.643 | 0.008 |
| Sex | Female (ref) |  |  |  |  |  |  |
|  | Male | 1.099 | 0.668 | 1.806 | 0.254 | 0.371 | 0.711 |
| Diagnosis | BVFTD (ref) |  |  |  |  |  |  |
|  | CBS | 2.297 | 0.940 | 5.610 | 0.456 | 1.825 | 0.068 |
|  | NFPPA | 0.923 | 0.365 | 2.333 | 0.473 | -0.169 | 0.865 |
|  | PSP | 4.481 | 1.809 | 11.099 | 0.463 | 3.241 | 0.001 |
|  | SVPPA | 0.411 | 0.136 | 1.249 | 0.566 | -1.568 | 0.117 |
| Severity | NfL | 1.002 | 0.999 | 1.004 | 0.001 | 1.509 | 0.131 |
|  | Symptom<br>Duration | 0.899 | 0.801 | 1.008 | 0.059 | -1.823 | 0.068 |
|  | ACE-R | 0.980 | 0.966 | 0.995 | 0.008 | -2.691 | 0.007 |

*Abbreviations: bvFTD=behavioural variant frontotemporal dementia; nfPPA=non-fluent primary progressive aphasia; svPPA=semantic variant primary progressive aphasia; PSP= progressive supranuclear palsy; CBS= corticobasal syndrome; HR=hazard ratio; CI= confidence interval; SE= standard error*

### Peripheral blood and brain PET markers of inflammation are correlated

Multivariate regression models on cytokine-derived component individual scores used regional values of [^11^C]PK11195 PET (marker of central inflammation) as well as age, sex and diagnosis as independent variables. The analysis of variance of the models identified levels of inflammation in the left frontal lobe (Estimate=15.51, t-value=1.76, F=4.78, p=0.036) and in the brainstem (Estimate=9.78, t-value=2.14, F=6.54, p=0.015) as statistically significant and positively associated predictors of cytokine-derived Component 1 individual scores. The other regional values, age, sex, and diagnosis were not statistically significant. After stepwise backward selection, the final model of multiple regression on cytokine-derived Component 1 (adjusted R2=0.188; P=0.005) included inflammation regional values in the left frontal lobe (Estimate=7.85, t-value=1.81, F=5.19, p=0.028) and brainstem (Estimate=9.02, t-value=2.61, F=6.79, p=0.0127) as predictors. Univariate associations between each significant predictor and individual scores of cytokine-derived Component 1 are described in Figure 3A, with frequentist and Bayesian Spearman’s correlation results. Exploratory correlation analyses were performed between regional [^11^C]PK11195 binding potentials across the whole brain and cytokine-derived Component 1 individual scores (Figure 3B). Significant positive correlations were found with the left middle frontal gyrus (rho=0.262), the left orbitofrontal cortex (rho=0.332), the left precentral gyrus (rho=0.420), the midbrain (rho=0.375) and pons (rho=0.324), and white matter of cerebellum (rho=0.314; p < 0.05).

**Figure 3.**
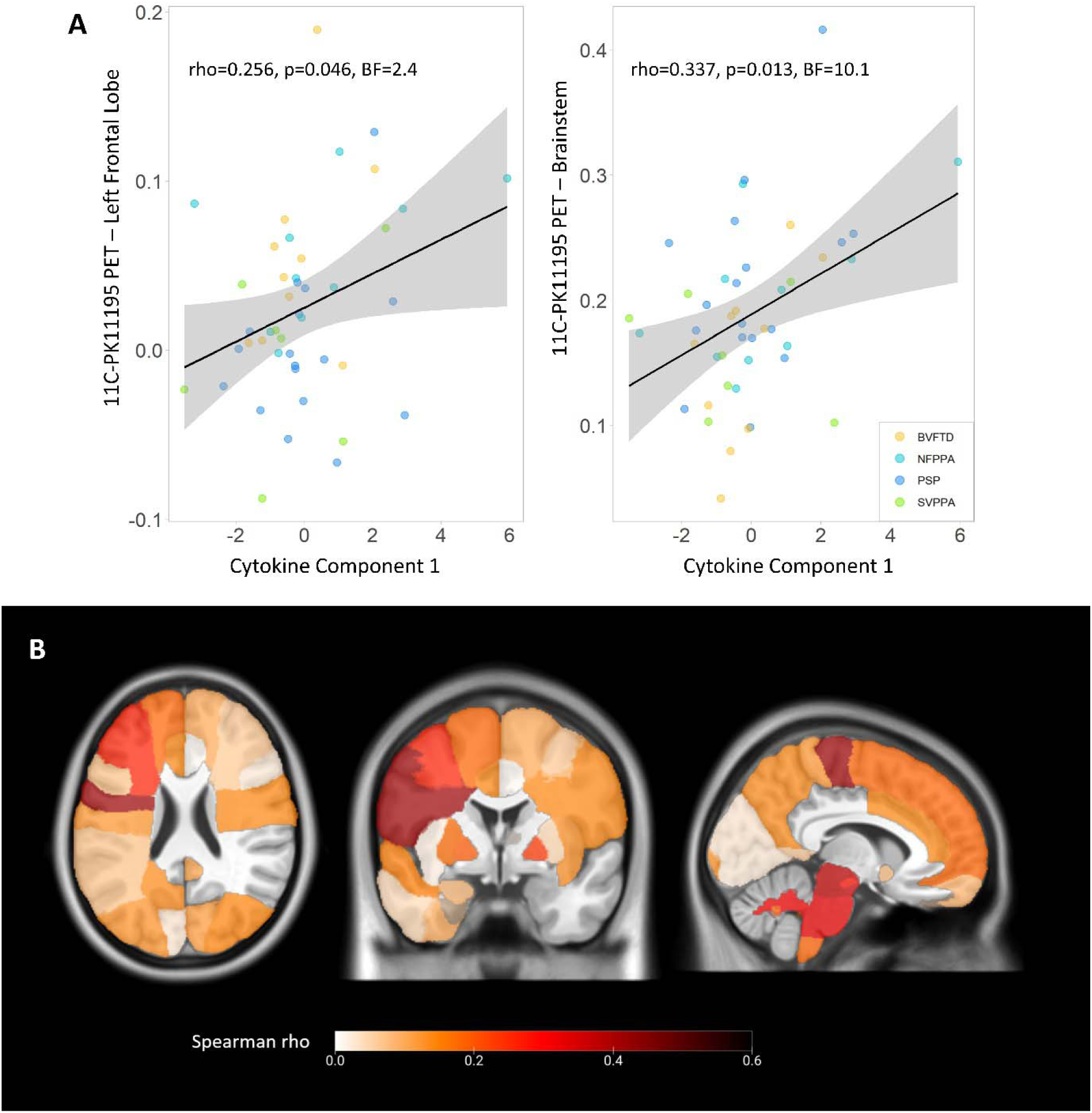
Associations between peripheral and central inflammation. Panel A: correlations between cytokine-derived Component 1 individual scores and [^11^C]PK11195 BPND in left frontal lobe and brainstem, which were identified as significantly associated with Component 1. Panel B: positive correlation coefficients (Spearman’s rho) from explorative regional analyses between cytokine-derived Component 1 individual scores and regional [^11^C]PK11195 binding potentials across the whole brain.

Finally, we used canonical correlation analysis (CCA) to test the association between cytokine-derived patterns and central brain inflammation in a fully data-driven approach. This revealed one significant component (all groups r=0.417, p=0.036). This component indicated a positive association between a pattern of cytokines loaded onto TNF-α, TNF-R1, M-CSF, IL-17A, IL-12, IP-10 and IL-6 (Figure 4A, similar to Component 1 in Figure 1A), and a pattern of microglial activation mainly weighted by PET signal in brainstem, frontal and parietal regions (Figure 4B). Higher individual scores in the cytokine-derived component were associated with higher individual scores in the PET-derived component, after correcting for age, sex and diagnosis (Estimate=0.41, t-value=3.19, F=23.7, p<0.0001, Figure 4C).

**Figure 4.**
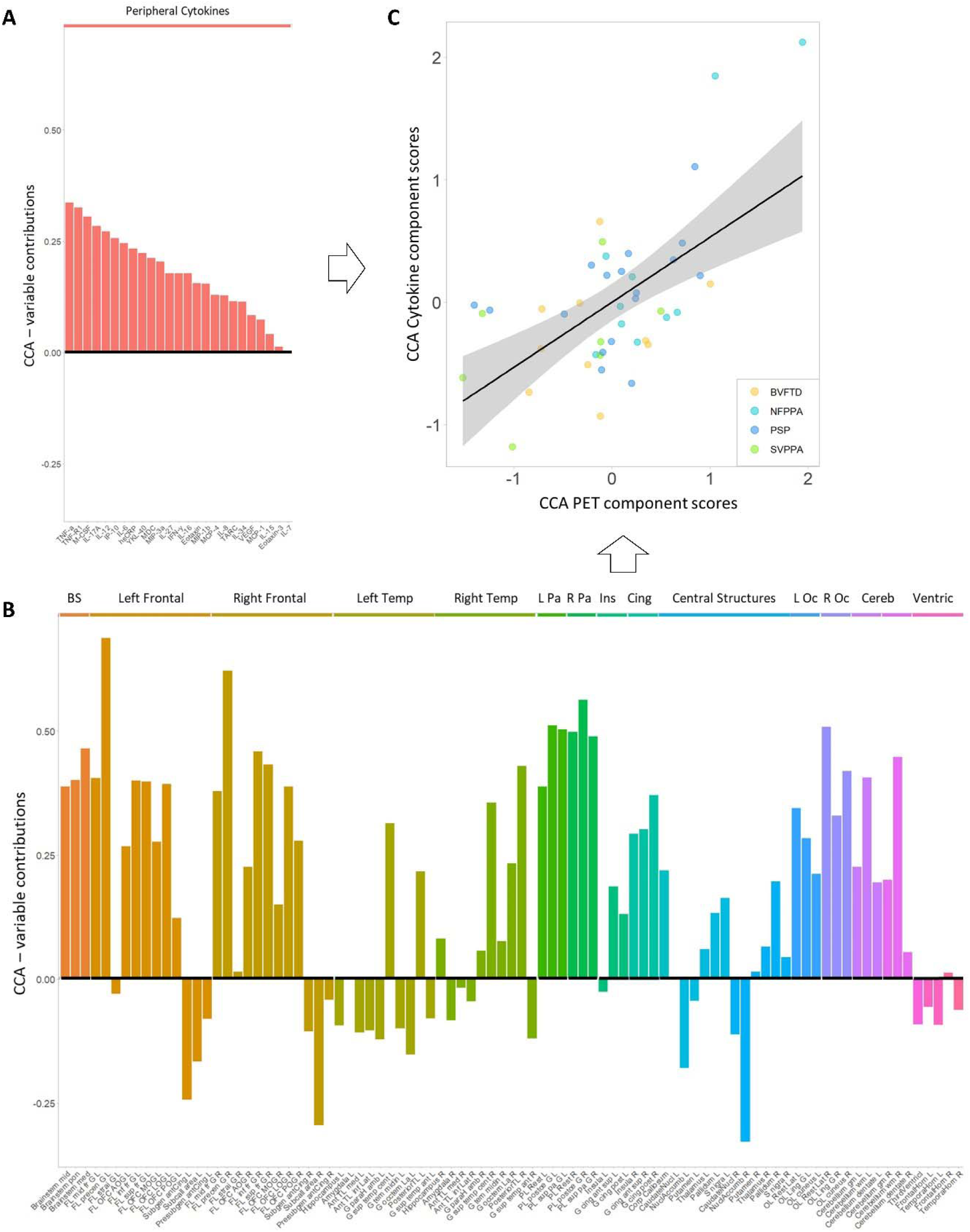
Comfirmatory Canonical Correlation Analysis (CCA) on cytokines and TSPO PER regional values. The bar graph showes contribution loadings for each variable to the first CCA component, which combines serum cytokines and [^11^C]PK11195 PET BPND regional values.

## Discussion

This study identified a clinically relevant profile of pro-inflammatory serum cytokines across FTLD-associated conditions, which differentiates patient groups from controls, and is expressed across all clinical syndromes. The inflammatory cytokine pattern was positively associated with higher levels of cerebral microglial activation on [^11^C]PK11195 TSPO PET imaging. The degree to which a patient expressed this peripheral inflammatory profile correlated with survival. We suggest that blood-based markers of inflammation could increase the scalability and access to neuroinflammatory assessment of people with dementia, to facilitate clinical trials and experimental medicine studies.

Recent advances in blood markers of *in vivo* inflammation in other forms of dementia ^28–30^ have raised the ambition for FTLD-related conditions. Although fluid markers confirmed abnormally high levels of inflammation in patients with FTLD-related conditions ^31–35^ (see ^7^ for review), they have been largely confined to cerebrospinal fluid (CSF), small case reports, limited panels and/or single markers. CSF markers can be highly sensitive, but unlike PET imaging they do not reveal the regional distribution of inflammation, despite being invasive and of limited repeatability and patient acceptability. Instead, there is the need for clinically relevant and mechanistically informative blood markers of neuroinflammation, to accelerate drug-discovery and trial participation. Here we investigated the covariance of serum cytokines to define inflammatory profiles with data-driven approaches. We validated the clinical relevance of these profiles testing for associations with markers of neurodegeneration (including NFL), brain inflammation (PET), and clinical progression (survival).

The most prominent inflammatory profile, identified by our approach with both principal component and canonical correlation analyses, was driven by pro-inflammatory cytokines, including TNF-α, TNF-R1, M-CSF, IL-17A, IL-12, IP-10 and IL-6. This multivariate (multi-cytokine) pattern supports and expand the results of more selective prior studies. TNF-α and its receptors regulate a large range of physiological functions, including immune reactions to infections, cell death induction, immune surveillance, and play a central role in initiating and regulating the cytokine cascade during an inflammatory response. Several studies have described elevated TNF-α levels in patients with Alzheimer’s disease (see metanalysis ^36,37^). When TNF-α signalling is inhibited both in patients with Alzheimer’s disease and transgenic mouse models, Alzheimer’s pathology is attenuated and cognition improves. Epidemiological studies suggest blocking TNF-α may reduce risk for Alzheimer’s disease in those with autoimmune conditions ^38,39^; while the upregulation of TNF-α leads to the exacerbation of pathology ^40^. However, studies on TNF-α in FTLD cohorts are limited. A recent study evaluated plasma levels of six cytokines in 39 patients with bvFTD; higher levels of TNF-α among other markers were associated with clinical severity and brain atrophy in frontal-limbic-striatal regions ^41^. The relevance of TNF-α in FTLD-related conditions was further confirmed by evidence from a large cohort of patients with pathogenic C9orf72, progranulin or MAPT variants, showing that higher levels of TNF-α at baseline are associated with faster clinical progression and more rapid brain atrophy rates longitudinally ^42^. In addition, plasma TNF-α improved the prediction of asymptomatic-to-symptomatic conversion, beyond plasma NfL alone ^42^. Elevated TNF-α levels were also previously observed in plasma of 129 patients with svPPA compared to neurologically healthy controls ^17^. Our study expands this series of blood-based marker studies, investigating a larger panel of inflammatory markers in patients with FTD, PPA, PSP and CBS, recruited in clinic- and epidemiologically based research studies. Collectively, our and previous results suggest that pro-inflammatory peripheral profiles, especially driven by TNFα, may contribute to FTLD disease progression and inform disease prognosis, both in sporadic and genetic forms of the disease.

Beyond TNF-α, previous studies on blood-based inflammation biomarkers in FTLD reported an increase of plasma IL-6 levels in 230 patients with FTD, irrespective of the clinical and genetic disease subtype ^43^, as well as increased serum levels of IL-6 in 14 carriers of GRN mutations ^44^. Studies on serum/plasma levels of M-CSF, IL-17A, IL-12, and IP-10 in patients with FTLD are limited or inconclusive. In contrast to previous studies, our approach combines a large panel of serum cytokines with data-driven methods to define inflammatory profiles, and includes patients from a large range of clinical syndromes and diseases related to FTLD. This approach may provide a comprehensive and complementary framework of inflammatory pathways and immune interactions than the application of single markers and hypothesis-driven analysis in single-disease cohorts.

The cytokine-derived proinflammatory profile (Component 1), in addition to differentiating patient groups from healthy controls, showed prognostic value. Individuals with higher scores in expressing this inflammatory profile were associated with subsequent faster decline and shorter time from blood sampling to death, over and above age, sex, and diagnosis, and indexes of disease baseline severity (NfL levels, symptom duration and cognitive performance). In particular, here we considered survival rate to test for the prognostic utility of inflammatory profiles, as the high clinical heterogeneity across the FTLD spectrum and measurement ceiling effects make difficult to capture clinical progression with a single cognitive test or questionnaire over time. In line with our results, genome-wide association studies implicated inflammatory pathways in the aetiology and progression of FTLD-related conditions. For example, associations were found between a common variation at the leucine-rich repeat kinase 2 (LRRK2) locus and survival from symptom onset to death in patients with PSP, which may be mediated by the effect of increased LRRK2 expression in microglia proinflammatory responses ^15^. Plasma NfL has been highlighted as promising marker for disease progression and as useful trial endpoint in patients with FTLD ^45–47^. Adding NfL levels to the survival model resulted in the cytokine-derived component retaining its predictive value. This suggests that combining biomarkers of neuronal damage and inflammation, such as TNF-α levels, may improve patient-specific prognostic estimations. This aligns with our previous studies showing that neuroinflammation levels, as measured with TSPO PET, provide additional predictive information on clinical and cognitive decline across the FTLD, over and above measures of brain atrophy, like structural MRI ^25,27^.

Importantly, we found that proinflammatory cytokine profiles (i.e. Component 1) identified in serum samples of patients with FTLD-related conditions are positively associated with higher levels of microglial activation in FTLD-specific brain regions. Although, blood-based quantification of inflammatory protein levels does not capture entirely the central inflammation processes happening in the brain, peripheral and central immune responses most likely engage in bidirectional interactive processes. Putative mechanistic linkages between peripheral and central immune interaction have been proposed: direct pathways by which peripheral immune cells may directly infiltrate the central nervous system; as well as indirect pathways by which systemic inflammation could drive a chronic modulation of microglial function ^48^. *Post mortem* studies reveal increased numbers of activated microglia and other neuroinflammatory responses in disease-specific regions that herald and topographically mirror neurodegeneration ^9,12,13,49^, including T cells infiltration in the brain and increased cytokine concentrations. TNF-α is synthesized by microglial cells following activation, as well as peripheral blood mononuclear cells, and can induce the activation of additional resting microglial cells or astrocytes. Along with TNF-α, interferon gamma (IFN-γ), which is a potent stimulator of other inflammatory cytokines, and YKL-40 also significantly contributed to the cytokine-derived Component 1. IFN-γ from both CD4+ and CD8+ T-cells promotes astrocyte proliferation and activation, resulting in exacerbated neuroinflammation. Increased IFN-γ expression is associated with T-cell brain infiltration and activated microglia with increased blood-brain barrier permeability ^50^. YKL-40 is also upregulated in several neurological disorders, and during neuroinflammatory processes, it is overexpressed in reactive astrocytes and microglial cells ^51^. The association between proinflammatory cytokine levels and regional microglial activation suggest that blood-based biomarkers of inflammation may be able to capture inflammatory processes that are relevant for central inflammation and brain changes. Further mechanistic studies are critically needed to clarify the crosstalk between peripheral and central immune systems in FTLD, and whether this interaction may represent a valid therapeutic target. Longitudinal clinical studies are also needed to clarify changes in the central-peripheral immune dynamics along the disease course.

Our study has limitations. Patients were included according to clinical diagnosis: post-mortem confirmation of pathology is available so far only in a small number of cases. This requires caution in the interpretation of likely underlying molecular pathology, especially in clinical syndromes with poor clinic-pathological correlation, such as cases of CBS ^52^ or two similarly prevalent alternative pathologies like bvFTD (Tau vs TDP43). Co-pathology may also occur, especially in older adults with age-related increase in the prevalence of Alzheimer pathology. Statistical power also needs consideration. We included a large sample size of the patient with FTLD, leading to well powered analyses. However, the lack of statistically significant differences between controls and patients with svPPA on individual scores of cytokine-derived Component 1 may be related to the relative smaller sample of the specific sub-cohort (n=20). Future studies on svPPA specifically with larger cohorts may be needed to identify statistically significant differences in cytokines levels, as per elevated plasma TNF-α levels described in Miller et al ^17^. CSF samples were available in a small group of patients (n<10), thus CSF-blood correlations were not achievable with meaningful power. However, we included TSPO PET as measure of brain inflammation to validate the relevance of the cytokine-derived proinflammatory pattern for central inflammatory processes. Blood-based inflammatory markers are not disease-specific and may capture comorbidities that are not directly linked to dementia. Co-occurring inflammatory conditions, or the use of specific anti-inflammatory medication were not systematically recorded. However, at the recruitment we excluded patients with co-morbid pro-inflammatory conditions, such as rheumatoid arthritis, inflammatory bowel disease, psoriasis or other autoimmune disorders and cancer. Our cohort was predominantly white/Caucasian reflecting the ethnicity distribution of the over 65-year-old population in the UK (94% white in the 2021 census), thus further studies are needed to generalise these results to other racial groups and to identify environmental, genetic and socioeconomical factors that might influence the immune responses in FTLD. Finally, the inflammatory markers were measured at a single time point, whilst longitudinal studies would be needed to test for disease-related dynamics of inflammation.

In conclusion, our data-driven approach identified a pro-inflammatory profile across each of the major FTLD-related conditions, which is positively associated with worse survival outcome and higher levels of brain microglial activation. Our results indicate the relevance of peripheral markers of inflammation to predict clinical progression in patients across the FTLD spectrum. Blood-based tests could greatly increase the scalability and access to neuroinflammatory assessment in dementia and experimental medicine studies. The combination of blood-based biomarkers for inflammation with makers for neurodegeneration (i.e. NfL) to evaluate patients at baseline may be a valuable approach to stratify patients into those likely to exhibit slow *versus* fast decline, improving cohort selection for clinical trials, and the interpretation of clinical outcomes.

## Methods

### Participants

Two hundred and fourteen (n=214) patients with clinical diagnoses associated with FTLD were recruited from specialist clinics at the Cambridge Centre for Frontotemporal Dementia and Cambridge Centre for Parkinson-plus. Exclusion criteria included recent or current acute infection, major concurrent psychiatric illness, other severe physical illness, or a history of other significant neurological illness. Twenty-nine healthy controls were recruited, with mini-mental state examination >26/30 with no acute physical illness, no cognitive complaints and independent in daily function and instrumental activities of daily living.

All participants underwent phlebotomy and a standard battery of cognitive tests and questionnaires, which included the revised version of Addenbrooke’s Cognitive Examination (ACE-R) ^53^, and the Revised Cambridge Behavioural Inventory (CBI-R). A sub-cohort of 44 patients (10 bvFTD, 10 nfvPPA, 7 svPPA, 17 PSP) underwent [^11^C]PK11195 PET as part of the NIMROD study (Neuroimaging of Inflammation in Memory and Related Other Disorders) ^54^.

Participants with mental capacity gave their written informed consent to take part in the study. For those who lacked capacity, their participation followed the consultee process in accordance with the UK law. The research protocols were approved by the National Research Ethics Service’s East of England Cambridge Central Committee, and the UK Administration of Radioactive Substances Advisory Committee.

### Blood sample collection and processing

Blood samples were obtained by venipuncture and collected in EDTA and serum gel tubes. Samples were centrifuged to isolate serum and plasma, aliquoted and stored at −70°C until further analyses. Serum cytokine analyses were carried out at the NIHR Cambridge Biomedical Research Centre, Core Biochemistry Assay Laboratory of Cambridge University Hospitals HNS Foundation Trust. The assays used the MesoScale Discovery (MSD) electrochemiluminescence immunoassay V-Plex Human Cytokine 36 plex panel and five additional cytokine assays: high sensitivity c-reactive protein (hsCRP, using Siemens Dimension EXL autoanalyser), tumour necrosis factor receptor 1 (TNF-R1) and interleukin-34 (IL-34) (using MSD R-plex assays), YKL-40 (chitinase-3-like protein 1) and colony stimulating factor 1 (using MSD R-plex assays). Dilutions were made in accordance with manufacturer recommendations. The MSD assays were performed in duplicate, with the mean taken for the purposes of analysis. The CRP analysis was performed in singleton. Further details of the cytokine assays and markers can be found in Supplementary Table 1.

In a sub-cohort of 186 patients (44 bvFTD, 29 nfvPPA, 20 svPPA, 44 PSP, 49 CBS), plasma samples were stored at −70°C for further analyses at the Clinical Neurochemistry Laboratory in Mölndal (Sweden). Plasma samples were thawed on wet ice, centrifuged at 500×□g for 5□min at 4°C. Calibrators (neat) and samples (plasma: 1:4 dilution) were measured in duplicates. The plasma assays measured were the Quanterix Simoa Human Neurology 4-Plex E assay (measuring Aβ40, Aβ42, GFAP and NfL, Quanterix, Billerica, MA) and the pTau217 ALZPath assay measuring p-tau217 of the human tau protein associated with Alzheimer’s disease, as previously described ^55^. Plasma samples were analysed at the same time using the same batch of reagents. A four-parameter logistic curve fit data reduction method was used to generate a calibration curve. Two control samples of known concentration of the protein of interest (high-control and low-control) were included as quality control.

### Imaging data acquisition and pre-processing

Structural MRI and [^11^C]PK11195 PET data were acquired and processed using previously described methods ^54^. Briefly, patients underwent a 3T MRI scan, followed by a dynamic [^11^C]PK11195 PET scan for 75 minutes at the Wolfson Brain Imaging Centre, University of Cambridge. MRI used Siemens Magnetom Tim Trio and Verio scanners (Siemens Healthineers, Erlangen, Germany) with an MPRAGE T1-weighted sequence, while PET used GE Advance and GE Discovery 690 PET/CT (GE Healthcare, Waukesha, USA) scanners. Time interval between blood sampling and PET scans was (mean ± standard deviation) 1.4 ± 2.8 months. Each T1 image was non-rigidly registered to the ICBM2009a template brain using ANTS (http://www.picsl.upenn.edu/ANTS/) and the inverse transform was applied to the Hammers atlas (resliced from MNI152 to ICBM2009a space) to bring the regions of interest to subject MRI space. The T1-weighted images were segmented into grey matter, white matter and cerebrospinal fluid (CSF) with SPM12 and used to determine regional grey matter, white matter and CSF volumes, and to calculate the total intracranial volume (grey matter + white matter + CSF) in each participant.

For each subject, the aligned dynamic PET image series for each scan was rigidly co-registered to the T1-weighted MRI image. Grey matter volumes and [^11^C]PK11195 non-displaceable binding potential (BP_ND_) for each tracer were calculated in cortical and subcortical ROIs using a modified version of the Hammers atlas. Prior to kinetic modelling, regional PET data were corrected for partial volume effects from CSF. Supervised cluster analysis was used to determine the reference tissue time-activity curve for [^11^C]PK11195 and BP_ND_ values were calculated in each ROI using a simplified reference tissue model with vascular binding correction ^56^.

### Statistical analyses

Statistical analyses used R 4.1.2 (R Core Team – see specific functions in the text), JASP and Matlab 2021b.

Age, years of education, and baseline cognitive/clinical scores were compared between groups with analysis of variance (ANOVA) tests, while sex was compared with the Chi-square test (see Table 1). For frequentist tests, p<0.05 after correction for multiple comparisons was considered significant (False Discovery Rate correction – FDR), uncorrected p<0.05 were described for explorative analyses. For Bayesian test, Bayesian Factor (BF) >3 indicates positive evidence for the alternative hypothesis and BF >10 indicates strong evidence for the alternative hypothesis; while BF<0.33 and BF < 0.1 indicate positive and strong evidence in favour of the null hypothesis respectively.

Sixteen of 41 cytokine markers were below the limit of detection for > 50% participants and thus excluded from further analyses. For the remaining 25 cytokine markers, values under the threshold of detectability were replaced with the variable-specific detection threshold. Serum cytokine concentrations were log10 transformed, and outliers were excluded after being defined as values over or below 5 standard deviations from the variable-specific mean (0.2% of total 6,075 cytokine values). This approach seeks to exclude extreme values due to methodological issues rather than pathophysiological changes. For further analyses, missing values generated by the exclusion of outliers were replaced by variable-specific means.

First, to test for dissimilarity between groups across all cytokines, values for each cytokine were averaged within group, creating a 25-cytokine vector for each group. Group-specific cytokine vectors entered a dissimilarity matrix computation analysis (R function *dist*, with the “Manhattan” method), which used the absolute distance between vectors to compute the distances between the rows of the data matrix. The resulting dissimilarity matrix was included in a hierarchical cluster analysis using the complete linkage (or furthest neighbour) method.

Second, cytokine marker values were included in a principal component analysis (PCA; R function *prcomp*), with the aim to reduce dimensionality and identify a limited number of *cytokine patterns* that best explain the data variance. We applied the Bartlett’s Test for Sphericity to test the null hypothesis that there was no collinearity between the variables included in PCA, and to verify that a data reduction technique like PCA can actually compress the data in a meaningful way. The number of components to be retained was defined by: (i) components with eigenvalues higher than 1 (Kaiser criterion); (ii) explained variance > 10%; (iii) scree plot review of the “break” or “elbow” (Cattell criterion).

Third, to test group differences in these cytokine principal components, individual scores of the resulting components were included in Kruskal–Wallis test with group as factor (R function *kruskal_test*). Post-hoc Dunn tests were applied to compare each diagnostic group to healthy controls (R function *dunn_test*). Associations of cytokine component individual scores with clinical outcomes, and plasma markers, were tested with linear regression models, including age, sex and diagnosis as covariates. We applied both frequentist and Bayesian analysis approaches to ensure inferential robustness, allowing us to quantify evidence in favour of the null hypothesis (of no group differences).

Fourth, we investigated the relationship between cytokine-derived component scores and survival. Survival data were available for 208 of the patients (141 deceased and 67 still alive; 49 bvFTD, 53 CBS, 31 nfPPA, 55 PSP, and 20 svPPA), at the census date 17^th^ October 2023. Survival analysis used Cox proportional hazards regression (R function *coxph*), including cytokine components scores and years from blood test to death as variables of interest, and age, sex and diagnosis group as covariates. A further explorative analysis was performed in a sub-group of 133 participants (77 deceased) to accommodate baseline severity: including baseline plasma NfL levels, symptoms duration (time interval between symptom onset and blood sampling), and ACE-R scores.

Fifth, we tested the association between cytokine-derived component individual scores and plasma levels of neurofilament light (NfL), Glial fibrillary acidic proteins (GFAP) and phosphorylated tau 217 (pTau217). We applied Spearman’s correlation and multiple regression models with each of the three plasma markers of interest as dependent variables, and cytokine-derived component individual scores, as well as age, sex and diagnosis as independent variables. Bayesian Spearman’s correlation (https://osf.io/gny35/) was secondarily applied to quantify evidence in favour of the null hypothesis (of no associations).

Finally, we tested the association between cytokine-derived component individual scores and central inflammation, quantified as regional [^11^C]PK11195 PET binding potential. Based on previous publications in patients with FTD ^20,25^ and PSP ^24,27^, regional [^11^C]PK11195 PET values were averaged within five hypothesis-driven regions of interest representing areas of peak neurodegenerative pathology: bilateral frontal and temporal lobes, and brainstem. A multiple regression model was fitted to examine the individual, as well as combined, ability to explain variance in cytokine-derived component individual scores using regional values of central inflammation as well as age, sex and diagnosis as independent variables. The model used stepwise backward selection (entry criterion α = 0.05 and elimination criterion α = 0.1) to identify the optimal model.

To confirm our results and the association between peripheral signatures of inflammation and microglial activation in the central system, beyond our hypothesized regions of interest, we examined the relationship between serum cytokines and whole-brain regional [^11^C]PK11195 PET values with a two-level analytical approach ^57–59^. In the first-level analysis, we assessed the multidimensional brain-cytokines relationships using permutation-based canonical correlation analysis (CCA). This analysis described the linear relationship between the two multivariate datasets ^60^ that are mapped to latent, common factors, or canonical variates, underlying these associations ^58^. Namely, dataset 1 consisted of the 25 cytokines in the previous analysis. Dataset 2 included the 89 Hammers-based regional [^11^C]PK11195 PET binding potentials. For this approach, instead of replacing missing cytokine values by variable-specific means, we imputed values under fully conditional specification, using the default settings of the multivariate imputation by chained equations (MICE) in R. Should the cytokine-derived component be similar to the one obtained with the first approach, this would ensure that results are not driven by a specific replacement approach. All variables were then standardized into z-scores before CCA. The CCA was permuted 5000 times to determine significance and ensure stability of the final components. Next, we tested whether the identified cytokine-relevant canonical variate of brain inflammation remained significant after correcting for age, sex and diagnosis, using a second-level linear regression analysis.

## Data availability

Anonymized processed data can be shared upon request to the corresponding or senior authors. Raw data may also be requested but are likely to be subject to a data transfer agreement with restrictions required to comply with participant consent and data protection regulations.

## Acknowledgments

This study was co-funded by the Dementias Platform UK and Medical Research Council (MC_UU_00030/14; MR/T033371/1); the Wellcome trust (220258); Race Against Dementia Alzheimer’s Research UK (ARUK-RADF2021A-010); the Cambridge University Centre for Parkinson-Plus (RG95450); the National Institute for Health Research (NIHR) Cambridge Biomedical Research Centre (NIHR203312: the views expressed are those of the authors and not necessarily those of the NIHR or the Department of Health and Social Care); the Progressive Supranuclear Palsy Association (PSPA2022/SMALL GRANTS002); the Addenbrookes Charitable Trust (ref 900380); Guarantors of Brain (G101149); Alzheimer’s Society (Grant Nr. 602). This work is licensed under a Creative Commons Attribution 4.0 International License. HZ is a Wallenberg Scholar and a Distinguished Professor at the Swedish Research Council supported by grants from the Swedish Research Council (#2023-00356; #2022-01018 and #2019-02397), the European Union’s Horizon Europe research and innovation programme under grant agreement No 101053962, Swedish State Support for Clinical Research (#ALFGBG-71320), the Alzheimer Drug Discovery Foundation (ADDF), USA (#201809-2016862), the AD Strategic Fund and the Alzheimer’s Association (#ADSF-21-831376-C, #ADSF-21-831381-C, #ADSF-21-831377-C, and #ADSF-24-1284328-C), the Bluefield Project, Cure Alzheimer’s Fund, the Olav Thon Foundation, the Erling-Persson Family Foundation, Stiftelsen för Gamla Tjänarinnor, Hjärnfonden, Sweden (#FO2022-0270), the European Union’s Horizon 2020 research and innovation programme under the Marie Skłodowska-Curie grant agreement No 860197 (MIRIADE), the European Union Joint Programme – Neurodegenerative Disease Research (JPND2021-00694), the National Institute for Health and Care Research University College London Hospitals Biomedical Research Centre, and the UK Dementia Research Institute at UCL (UKDRI-1003). We thank our participant volunteers for their participation in this study, thank the National Institute for Health Research (NIHR) Cambridge BioResource centre and staff, and the research nurses for their contribution, and the East Anglia Dementias and Neurodegenerative Diseases Research Network (DeNDRoN) for help with subject recruitment. In particular, we thank Alicia Wilcox, Carolyn Timberlake, David Whiteside, Hugo Paula, Ian Coyle-Gilchrist, Jacqueline Young, Julie Wiggins, Karalyn Patterson, Katherine Stockton, Luca Passamonti, Lucy Bowns, Michelle Naessens, Matthew Rouse, Negin Holland, Patricia Vázquez Rodríguez, Robert Durcan and Win Li for their assistance in the study, including recruitment and phlebotomy. For the purpose of open access, the authors have applied a Creative Commons Attribution (CC BY) license to any Author Accepted Manuscript version arising from this submission.

