## Supplementary Material for "Peripheral inflammatory markers relate to central inflammation and survival in syndromes associated with frontotemporal lobar degeneration"

**Supplementary Table 1. Details of Cytokine Assays.**

| **Cytokine** | **MSD Panel** | **Unit** | **LLOD** | **Source of LLOD** | **Included in analysis** |
| --- | --- | --- | --- | --- | --- |
| IFN-γ | MSD V-plex Proinflammatory Panel 1 | pg/ml | 2.12 | Calculated by MSD WorkBench software as part of the data interpretation from the analysis for this study | 1 |
| IL-10 | MSD V-plex Proinflammatory Panel 1 | pg/ml | 0.24 |  | 0 |
| IL-12p70 | MSD V-plex Proinflammatory Panel 1 | pg/ml | 0.26 |  | 0 |
| IL-13 | MSD V-plex Proinflammatory Panel 1 | pg/ml | 1.96 |  | 0 |
| IL-1β | MSD V-plex Proinflammatory Panel 1 | pg/ml | 1.4 |  | 0 |
| IL-2 | MSD V-plex Proinflammatory Panel 1 | pg/ml | 0.47 |  | 0 |
| IL-4 | MSD V-plex Proinflammatory Panel 1 | pg/ml | 0.07 |  | 0 |
| IL-6 | MSD V-plex Proinflammatory Panel 1 | pg/ml | 0.38 |  | 1 |
| IL-8 | MSD V-plex Proinflammatory Panel 1 | pg/ml | 0.26 |  | 1 |
| TNF-α | MSD V-plex Proinflammatory Panel 1 | pg/ml | 0.58 |  | 1 |
| Eotaxin | MSD V-plex Chemokine Panel 1 | pg/ml | 60 |  | 1 |
| Eotaxin-3 | MSD V-plex Chemokine Panel 1 | pg/ml | 9.87 |  | 1 |
| IP-10 | MSD V-plex Chemokine Panel 1 | pg/ml | 1.28 |  | 1 |
| MCP-1 | MSD V-plex Chemokine Panel 1 | pg/ml | 0.88 |  | 1 |
| MCP-4 | MSD V-plex Chemokine Panel 1 | pg/ml | 20.51 |  | 1 |
| MDC | MSD V-plex Chemokine Panel 1 | pg/ml | 29.12 |  | 1 |
| MIP-1α | MSD V-plex Chemokine Panel 1 | pg/ml | 25.01 |  | 0 |
| MIP-1β | MSD V-plex Chemokine Panel 1 | pg/ml | 3.48 |  | 1 |
| TARC | MSD V-plex Chemokine Panel 1 | pg/ml | 11.8 |  | 1 |
| GM-CSF | MSD V-plex Cytokine Panel 1 | pg/ml | 0.67 |  | 0 |
| IL-1α | MSD V-plex Cytokine Panel 1 | pg/ml | 0.82 |  | 0 |
| IL-12 | MSD V-plex Cytokine Panel 1 | pg/ml | 1.2 |  | 1 |
| IL-15 | MSD V-plex Cytokine Panel 1 | pg/ml | 0.42 |  | 1 |
| IL-16 | MSD V-plex Cytokine Panel 1 | pg/ml | 5.7 |  | 1 |
| IL-17A | MSD V-plex Cytokine Panel 1 | pg/ml | 2.6 |  | 1 |
| IL-5 | MSD V-plex Cytokine Panel 1 | pg/ml | 0.5 |  | 0 |
| IL-7 | MSD V-plex Cytokine Panel 1 | pg/ml | 0.94 |  | 1 |
| TNF-β | MSD V-plex Cytokine Panel 1 | pg/ml | 0.48 |  | 0 |
| VEGF | MSD V-plex Cytokine Panel 1 | pg/ml | 0.78 |  | 1 |
| IL- 17A | MSD V-plex TH17 Panel 1 | pg/ml | 14.62 |  | 0 |
| IL-21 | MSD V-plex TH17 Panel 1 | pg/ml | 19 |  | 0 |
| IL-31 | MSD V-plex TH17 Panel 1 | pg/ml | 0.8 |  | 0 |
| IL-27 | MSD V-plex TH17 Panel 1 | pg/ml | 66.96 |  | 1 |
| IL-23 | MSD V-plex TH17 Panel 1 | pg/ml | 13.04 |  | 0 |
| IL-22 | MSD V-plex TH17 Panel 1 | pg/ml | 1.35 |  | 0 |
| MIP-3α | MSD V-plex TH17 Panel 1 | pg/ml | 4.58 |  | 1 |
| IL-34 | MSD R-plex | pg/ml | 1.34 |  | 1 |
| TNF-R1 | MSD R-plex | pg/ml | 3.8 |  | 1 |
| M-CSF | MSD U-plex | pg/ml | 0.76 |  | 1 |
| YKL-40 | MSD U-plex | pg/ml | 2186 |  | 1 |
| hsCRP | Dimension | mg/L | 0.1 | Manufacturer | 1 |

*Abbreviations: LLOD=Lower Limit of Detection; MSD= MesoScale Discovery*

**Supplementary Table 2. Dunn’s Multiple Comparison post hoc test following the Kruskal-Wallis test on cytokine-derived Component 1.**

| **Group 1** | **Group 2** | **n1** | **n2** | **statistic** | **p** | **p FDR** |
| --- | --- | --- | --- | --- | --- | --- |
| HC | bvFTD | 29 | 52 | 3.119 | 0.002* | 0.019* |
| HC | svPPA | 29 | 20 | 1.715 | 0.086 | 0.259 |
| HC | nfPPA | 29 | 31 | 3.020 | 0.003* | 0.019* |
| HC | PSP | 29 | 58 | 2.316 | 0.021* | 0.077 |
| HC | CBS | 29 | 53 | 2.784 | 0.005* | 0.027* |
| bvFTD | svPPA | 52 | 20 | -0.852 | 0.394 | 0.657 |
| bvFTD | nfPPA | 52 | 31 | 0.252 | 0.801 | 0.858 |
| bvFTD | PSP | 52 | 58 | -1.028 | 0.304 | 0.612 |
| bvFTD | CBS | 52 | 53 | -0.409 | 0.683 | 0.788 |
| svPPA | nfPPA | 20 | 31 | 0.981 | 0.326 | 0.612 |
| svPPA | PSP | 20 | 58 | 0.108 | 0.914 | 0.914 |
| svPPA | CBS | 20 | 53 | 0.550 | 0.582 | 0.727 |
| nfPPA | PSP | 31 | 58 | -1.139 | 0.255 | 0.612 |
| nfPPA | CBS | 31 | 53 | -0.606 | 0.544 | 0.727 |
| PSP | CBS | 58 | 53 | 0.613 | 0.540 | 0.727 |

*Abbreviations: bvFTD=behavioural variant frontotemporal dementia; nfPPA=non-fluent primary progressive aphasia; svPPA=semantic variant primary progressive aphasia; PSP=progressive supranuclear palsy; CBS= corticobasal syndrome; FDR=False Discovery Rate correction*

**Supplementary Table 3. Loadings from cytokine variables (red) and regional TSPO PET values (yellow) on the first CCA component.**

| **Category** | **Variable** | **Loadings (contribution to CCA Component)** |
| --- | --- | --- |
| Cytokines | TNF-a | 0.337 |
| Cytokines | TNF-R1 | 0.325 |
| Cytokines | M-CSF | 0.305 |
| Cytokines | IL-17A | 0.284 |
| Cytokines | IL-12 | 0.271 |
| Cytokines | IP-10 | 0.257 |
| Cytokines | IL-6 | 0.246 |
| Cytokines | hsCRP | 0.233 |
| Cytokines | YKL-40 | 0.224 |
| Cytokines | MDC | 0.212 |
| Cytokines | MIP-3a | 0.204 |
| Cytokines | IL-27 | 0.178 |
| Cytokines | IFN-y | 0.178 |
| Cytokines | IL-16 | 0.178 |
| Cytokines | Eotaxin | 0.155 |
| Cytokines | MIP-1b | 0.155 |
| Cytokines | MCP-4 | 0.129 |
| Cytokines | IL-8 | 0.129 |
| Cytokines | TARC | 0.115 |
| Cytokines | IL-34 | 0.114 |
| Cytokines | VEGF | 0.084 |
| Cytokines | MCP-1 | 0.073 |
| Cytokines | IL-15 | 0.041 |
| Cytokines | Eotaxin-3 | 0.012 |
| Cytokines | IL-7 | -0.001 |
| TSPO PET | Brainstem mid | 0.387 |
| TSPO PET | Brainstem pon | 0.400 |
| TSPO PET | Brainstem med | 0.464 |
| TSPO PET | FL mid fr G L | 0.405 |
| TSPO PET | FL precen G L | 0.686 |
| TSPO PET | FL strai G L | -0.030 |
| TSPO PET | FL OFC AOG L | 0.266 |
| TSPO PET | FL inf fr G L | 0.400 |
| TSPO PET | FL sup fr G L | 0.397 |
| TSPO PET | FL OFC MOG L | 0.275 |
| TSPO PET | FL OFC LOG L | 0.393 |
| TSPO PET | FL OFC POG L | 0.122 |
| TSPO PET | Subgen antCing L | -0.244 |
| TSPO PET | Subcall area L | -0.167 |
| TSPO PET | Presubgen antCing L | -0.082 |
| TSPO PET | FL mid fr G R | 0.378 |
| TSPO PET | FL precen G R | 0.621 |
| TSPO PET | FL strai G R | 0.014 |
| TSPO PET | FL OFC AOG R | 0.225 |
| TSPO PET | FL inf fr G R | 0.458 |
| TSPO PET | FL sup fr G R | 0.432 |
| TSPO PET | FL OFC MOG R | 0.149 |
| TSPO PET | FL OFC LOG R | 0.388 |
| TSPO PET | FL OFC POG R | 0.277 |
| TSPO PET | Subgen antCing R | -0.107 |
| TSPO PET | Subcall area R | -0.296 |
| TSPO PET | Presubgen antCing R | -0.043 |
| TSPO PET | Hippocampus L | -0.094 |
| TSPO PET | Amygdala L | 0.004 |
| TSPO PET | Ant TL med L | -0.109 |
| TSPO PET | Ant TL inf Lat L | -0.104 |
| TSPO PET | G paraH amb L | -0.123 |
| TSPO PET | G sup temp cent L | 0.314 |
| TSPO PET | G tem midin L | -0.101 |
| TSPO PET | G occtem La L | -0.154 |
| TSPO PET | PosteriorTL L | 0.216 |
| TSPO PET | G sup temp ant L | -0.080 |
| TSPO PET | Hippocampus R | 0.081 |
| TSPO PET | Amygdala R | -0.085 |
| TSPO PET | Ant TL med R | -0.019 |
| TSPO PET | Ant TL inf Lat R | -0.046 |
| TSPO PET | G paraH amb R | 0.057 |
| TSPO PET | G sup temp cent R | 0.355 |
| TSPO PET | G tem midin R | 0.076 |
| TSPO PET | G occtem La R | 0.232 |
| TSPO PET | PosteriorTL R | 0.428 |
| TSPO PET | G sup temp ant R | -0.121 |
| TSPO PET | PL Rest L | 0.387 |
| TSPO PET | PL postce G L | 0.511 |
| TSPO PET | PL sup pa G L | 0.503 |
| TSPO PET | PL Rest R | 0.497 |
| TSPO PET | PL postce G R | 0.563 |
| TSPO PET | PL sup pa G R | 0.488 |
| TSPO PET | Insula L | -0.026 |
| TSPO PET | G cing ant sup L | 0.186 |
| TSPO PET | G cing post L | 0.130 |
| TSPO PET | Insula R | 0.292 |
| TSPO PET | G cing ant sup R | 0.301 |
| TSPO PET | G cing post R | 0.370 |
| TSPO PET | Corp Callosum | 0.218 |
| TSPO PET | CaudateNucl L | 0.004 |
| TSPO PET | NuclAccumb L | -0.180 |
| TSPO PET | Putamen L | -0.045 |
| TSPO PET | Thalamus L | 0.059 |
| TSPO PET | Pallidum L | 0.132 |
| TSPO PET | S nigra L | 0.163 |
| TSPO PET | CaudateNucl R | -0.113 |
| TSPO PET | NuclAccumb R | -0.329 |
| TSPO PET | Putamen R | 0.014 |
| TSPO PET | Thalamus R | 0.064 |
| TSPO PET | Pallidum R | 0.196 |
| TSPO PET | S nigra R | 0.043 |
| TSPO PET | OL Rest Lat L | 0.343 |
| TSPO PET | OL Ling G L | 0.283 |
| TSPO PET | OL cuneus L | 0.211 |
| TSPO PET | OL Rest Lat R | 0.507 |
| TSPO PET | OL Ling G R | 0.329 |
| TSPO PET | OL cuneus R | 0.418 |
| TSPO PET | Cerebellum gm L | 0.225 |
| TSPO PET | Cerebellum wm L | 0.405 |
| TSPO PET | Cerebellum dentate L | 0.194 |
| TSPO PET | Cerebellum gm R | 0.199 |
| TSPO PET | Cerebellum wm R | 0.447 |
| TSPO PET | Cerebellum dentate R | 0.053 |
| TSPO PET | Third Ventricl | -0.092 |
| TSPO PET | Frontal Horn L | -0.057 |
| TSPO PET | Tempora Horn L | -0.094 |
| TSPO PET | Frontal Horn R | 0.012 |
| TSPO PET | Tempora Horn R | -0.063 |

**Supplementary Figure 1. Dissimilarity between groups across all cytokines.** Darker colours and larger values indicate greater distance between groups, while lighter colours indicate relative similarity.

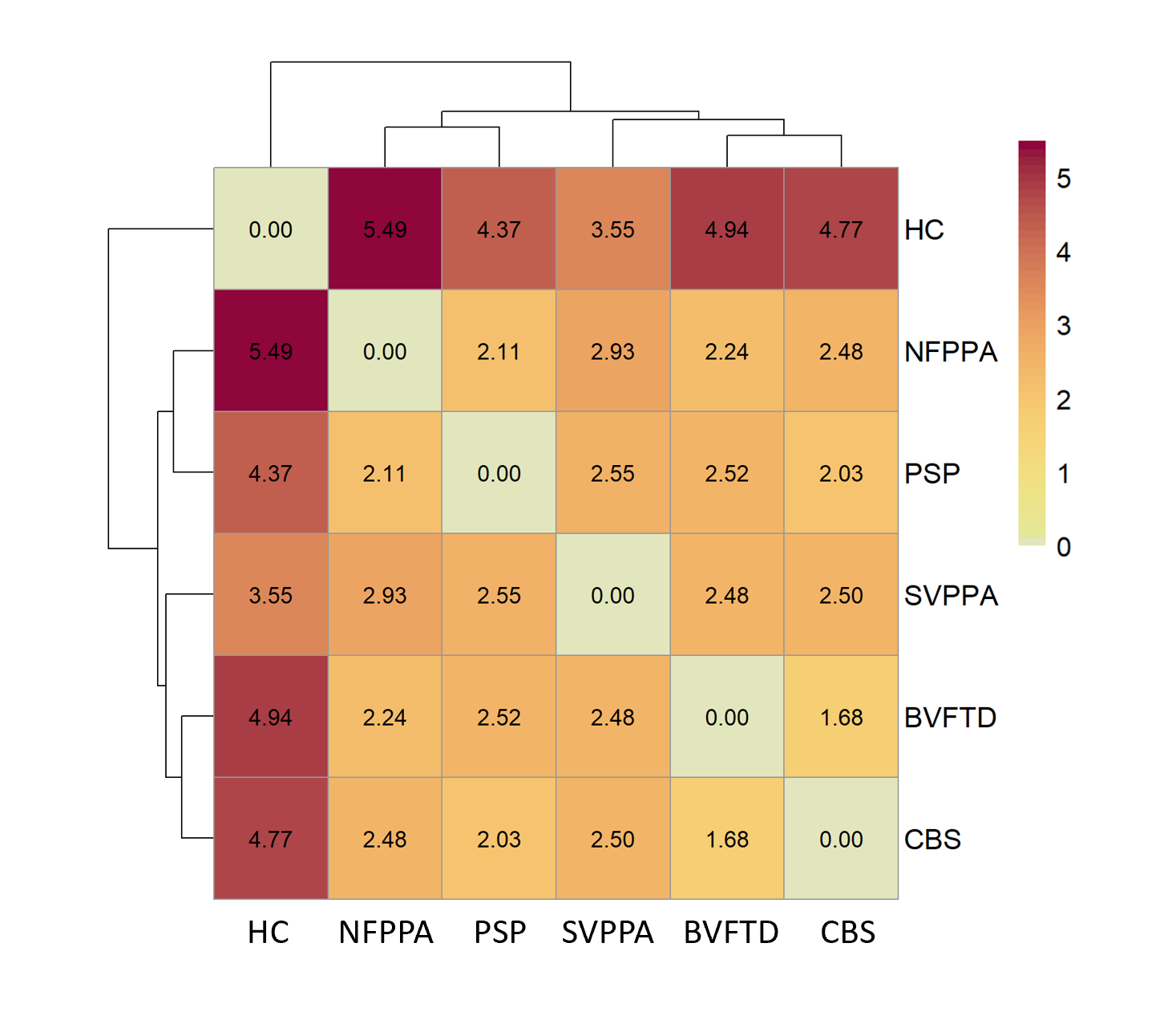

**Supplementary Figure 2. Associations between individual loadings of cytokine-derived Component 1 and plasma marker levels.**

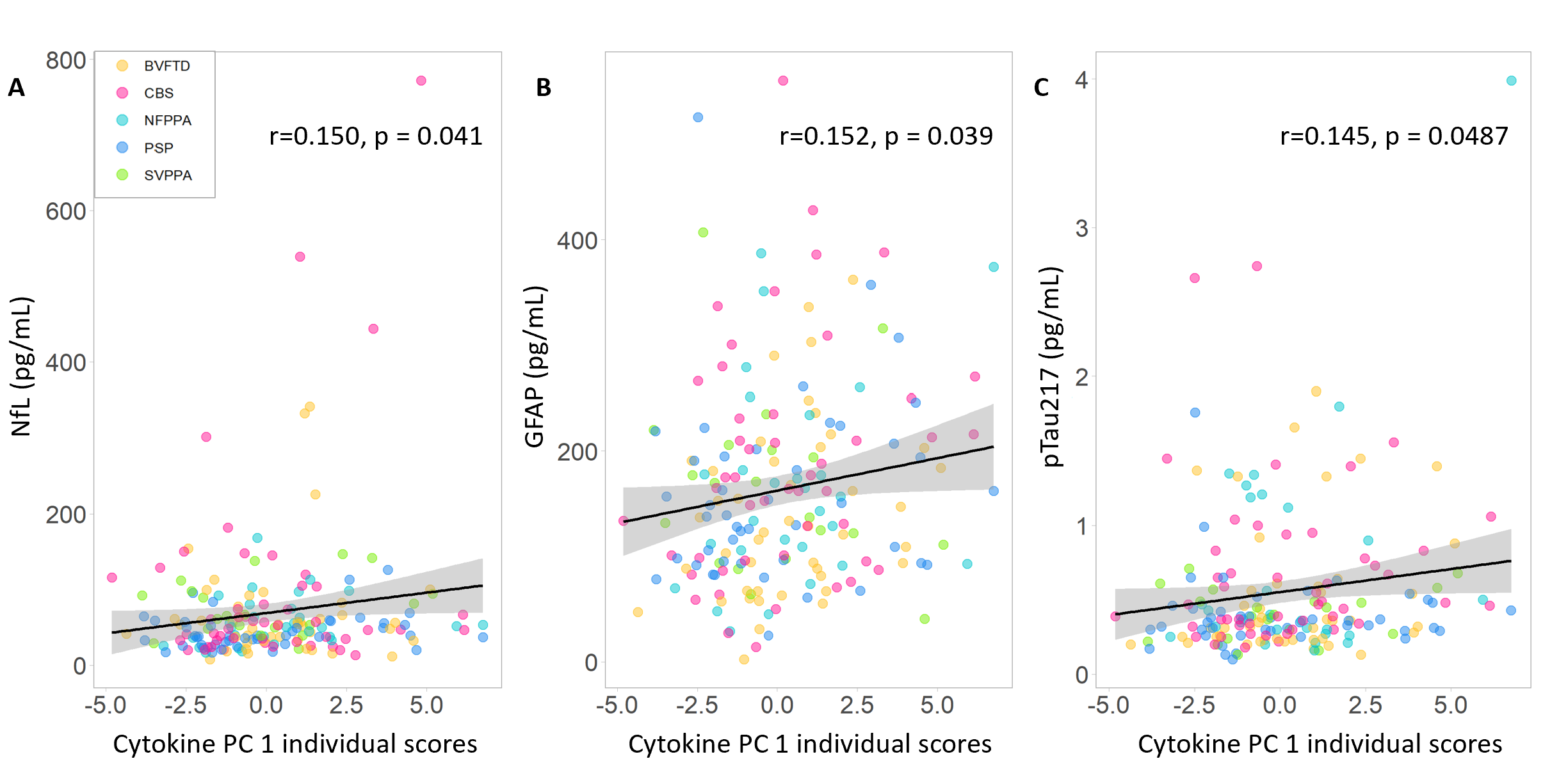
